# Multimodal MRI Reveals Consistent Basal Ganglia and Limbic System Alterations in COVID-19 Survivors

**DOI:** 10.1101/2025.06.20.25329994

**Authors:** Sapna S Mishra, Caterina A Pedersini, Rohit Misra, Preeti Yadav, Rakibul Hafiz, Bas Rokers, Bharat Biswal, Tapan K. Gandhi

## Abstract

The long-term impact of COVID-19 on the brain is multifaceted, encompassing structural and functional disruptions. A cohesive theory of the underlying mechanisms of the Post-COVID Syndrome (PCS) remains unknown, primarily due to high variability in findings across independent studies. Here, we present a multimodal, cross-sectional MRI analysis of brain morphology (T1-MRI), tissue microstructure (diffusion-MRI), functional connectivity (functional-MRI), and cerebral blood flow (Arterial Spin Labeling MRI) in COVID-Recovered Patients (CRPs, N=76) and Healthy Controls (HCs, N=51). Although the global brain volumes did not differ between the two groups, CRPs showed focal atrophy in the right basal ganglia and limbic structures, along with cortical thinning in paralimbic regions (prefrontal cortex, insula) (p<0.05). Diffusion MRI analysis revealed reduced fractional anisotropy and elevated radial diffusivity in the uncinate fasciculus and cingulum. No differences were observed in resting-state functional connectivity (RSFC) and cerebral blood flow between HCs and CRPs (p>0.05). We further investigated the effect of infection severity by stratifying the CRPs into hospitalized (HP; N = 21) and non-hospitalized (NHP; N = 46) groups. The microstructural damage was linked to infection severity, more pronounced in the HPs (p<0.05). In HPs, RSFC was diminished between components of the default mode network and the insula and caudate as compared to HCs and NHPs (p<0.05). Results suggest COVID-19 is associated with selective structural and functional alterations in basal ganglia–limbic–cortical circuits, with stronger effects in severe cases. Our findings are in line with common prevalent behavioral symptoms such as fatigue, memory impairment, attentional deficits, and insomnia. This study suggests that localized microstructural neuroinflammatory mechanisms contribute to post-COVID neurological symptoms and offers potential imaging biomarkers for targeted therapies and monitoring recovery.

## 1. Introduction

Coronavirus Disease 2019 (COVID-19) primarily manifests as a respiratory infection, characterized by common symptoms such as fever, cough, and fatigue [1]. Since its emergence in December 2019, COVID-19 precipitated a global pandemic with profound health, economic, and societal consequences. Healthcare systems worldwide were overwhelmed, economic activities disrupted, and daily life profoundly altered through public health interventions such as mask mandates, social distancing, and widespread vaccination campaigns. Although the respiratory symptoms of COVID-19 typically resolve within weeks, mounting evidence indicates that a substantial proportion of survivors experience persistent cognitive and behavioral symptoms even after virological recovery [2]. Reports of enduring “brain fog”, memory deficits, inattention, cognitive fatigue, and emotional disturbances have continued to surface months to years after the initial infection [2, 3]. These long-term manifestations, often grouped under the umbrella of Post-COVID Syndrome (PCS) or Long COVID, have raised major public health concerns due to their impact on survivors’ quality of life and functional independence.

A growing body of literature underscores the diversity and prevalence of these sequelae. Meta-analyses indicate that approximately 32% of COVID-19 survivors report significant fatigue 12 weeks after recovery [4], while about 22% exhibit measurable cognitive impairment. Sleep disturbances (27.4%), fatigue (24.4%), cognitive deficits (20.2%), anxiety (19.1%), and post-traumatic stress (15.7%) have emerged as the most common persistent symptoms in post-COVID cohorts [2]. Furthermore, cognitive performance studies have shown that COVID-19 survivors display significantly lower cognitive function compared to uninfected controls [5].

Neuroimaging has played a critical role in delineating the structural, microstructural, and functional brain alterations associated with COVID-19 recovery. Early studies on structural MRI revealed variable tissue changes. Niu *et al*. reported increased grey matter volumes in the posterior cingulate cortex and isthmus regions [6], whereas Douaud *et al*., analyzing UK Biobank data, found cortical thinning in the orbitofrontal cortex and parahippocampal gyrus following COVID-19 infection [7]. Alongside grey matter changes, diffusion-weighted MRI studies have uncovered widespread white matter abnormalities. Increased mean diffusivity (MD) and reduced fractional anisotropy (FA) were observed globally in COVID survivors [8], while Diez-Cirarda *et al*. reported tract-specific reductions in MD within the uncinate fasciculus (UF), corpus callosum, forceps minor, and fronto-occipital fasciculus (FOF) [9].

Functional MRI studies have further demonstrated that COVID-19 is associated with altered intrinsic brain connectivity. Leitner *et al*. identified impaired functional connectivity (FC) between the thalamus and cortical regions, including the precentral gyrus, supramarginal gyrus, and anterior cingulate cortex [10]. Reductions in FC within canonical networks such as the default mode network (DMN), dorsal attention network, and somatomotor network have also been reported in COVID-19 recovered patients [11, 12], suggesting that functional network changes may be linked to persistent cognitive and emotional deficits. Complementary studies using arterial spin labeling MRI have suggested cerebral blood flow (CBF) alterations in COVID-19 survivors. Kim *et al*. identified CBF reductions in the thalamus, orbitofrontal cortex, and basal ganglia [13], whereas Qin *et al*. reported decreased CBF in the superior medial frontal gyrus and insula [14]. These vascular findings suggest that microvascular dysfunction may be associated with pathogenesis of post-COVID neurological symptoms.

Taken together, existing studies highlight that COVID-19’s neurological impact is multifaceted, reflected in vascular, structural, and functional disruptions. However, a large section of post-COVID studies focuses on these aspects in isolation. Diversity in their findings obstructs attempts to form a cohesive theory of the mechanisms behind PCS and to identify the key brain regions or systems that are targeted by PCS. Multimodal neuroimaging approaches are, therefore, essential for the comprehensive characterization of these sequelae, and large-scale, well-powered investigations are needed to form an integrated understanding of PCS pathophysiology. Addressing this need, here we present a multimodal, cross-sectional MRI analysis of brain morphology, tissue microstructure, functional connectivity, and cerebral blood flow in COVID-19 survivors compared to healthy controls. Conducted in India, this large-scale study utilized T1-weighted MRI, diffusion-weighted MRI (dMRI), resting-state functional MRI (rs-fMRI), and ASL perfusion imaging. We first contrast COVID-19 Recovered Patients (CRPs) and Healthy Controls (HCs) across all modalities. We then stratify CRPs based on hospitalization status to explore the effects of disease severity. Finally, we synthesize findings across imaging techniques to propose mechanistic pathways linking brain injury with post-COVID symptomatology. Through this comprehensive investigation, we aim to contribute novel insights into the neural substrates of Long COVID.

## 2. Methods

### 2.1. Subject Recruitment

This cross-sectional study is based on two cohorts of subjects: (i) COVID-Recovered Patients (CRPs), and (ii) Healthy Controls (HCs). The recruitment of CRPs was done using a database of 2358 COVID-19 patients who were treated at a local hospital. Among these patients, 22% needed continuous positive airway pressure (CPAP), 40% received oxygen therapy, and 14% required intubation. After two weeks of testing PCR negative, 100 hospitalized patients from this database were contacted for this study. In addition, 126 CRPs who did not require hospitalisation were also contacted.

The inclusion criteria for the CRP cohort were (i) age above 18 years, (ii) diagnosis of COVID-19 (RT-PCR positive), and (iii) subsequent proof of recovery from infection (RT-PCR negative) within six months before the scan; Similarly, inclusion criteria for the HC cohort were, (i) age above 18 years, and (ii) no history of COVID-19 infection as on scan date (symptomatic subjects required a negative RT-PCR report to confirm). For both cohorts, exclusion criteria were a history of (i) neuropsychiatric or neurological disorders, (ii) brain surgery/trauma, or (iii) underwent ventilation during the infection (for CRPs). In total, we collected MRI scans of 76 CRPs (32.38 ± 11.52, 19 Female) and 51 HCs (31.98 ± 8.97 years, 12 Female). Later, certain MRIs had to be excluded from specific analyses due to poor image quality (details discussed in the respective section). Additionally, 67 CRPs (31.12 ± 10.78 years, 16 Female) consented to sharing their treatment history and 59 CRPs (30.85 ± 10.52 years, 15 Female) also reported the post-COVID symptoms experienced by them after a negative RT-PCR test.

Data collection occurred under the purview of the Indian Institute of Technology Delhi, and all imaging procedures were conducted at Mahajan Imaging Center, New Delhi in accordance with the Institute Review Board (IRB) regulations. The pilot study was approved by the ethics committee of the Mahajan Imaging Center, and the entire study was approved by the Institute Ethics Committee, IIT Delhi. All subjects provided informed consent before any behavioural or physical data was collected.

### 2.2. Classification Based on Infection Severity

Among the 76 CRPs included in this study, 67 consented to share their treatment history. We used the information from their treatment history to categorize them as Non-Hospitalized Patients or Hospitalized Patients, depending on whether they needed to be hospitalized during the treatment of their COVID-19 infection. In this study, we interpret the hospitalisation of a COVID-19 patient as a mark of severe infection. Subjects categorised as HPs needed procedures like CPAP, oxygen therapy, or continuous monitoring during their treatment. On the other hand, the COVID-19 symptoms of NHPs included fever, cough, loss of taste, loss of smell, and weakness, and they did not require hospitalization for treatment. Based on these criteria, 21 (39.48 ± 11.95 years, 6F) subjects were categorised as Hospitalized Patients (HPs) and the remaining 46 (27.30 ± 7.72 years, 10F) as Non-Hospitalized Patients (NHPs). The NHP and HP groups were used to test the relationship of MRI-derived measures with infection severity.

### 2.3. Magnetic Resonance Imaging

T1-weighted images were collected using a 3T GE Discovery MR 750w scanner, employing the fast BRAVO sequence in 3D imaging mode with a 32-channel head coil. Imaging parameters were: 12 Flip angle, 450*ms* inversion time (TI), 256 *mm* × 256 *mm* Field of View (FOV), 1.00 *mm* slice thickness, 152 sagittal slices, and voxels with 1 *mm* isotropic resolution.

dMRI scans were acquired with a spin-echo echo-planar imaging (EPI) sequence with the following parameters: 16000 *ms* Repetition Time (TR), 76 *ms* Echo time (TE), 256 × 256 matrix, 262 *mm* × 263 *mm* FOV, 2 *mm* slice thickness, 1 × 1× 2 *mm*^3^ voxel resolution. The sequence used 30 diffusion encoding directions with anterior-posterior phase encoding and *b* = 1000 *s/mm*^2^ for each direction. Four T2 volumes were also acquired with no diffusion encoding (*b* = 0 *s/mm*^2^).

The rs-fMRI scans were acquired using gradient echo planar imaging (EPI) with *TR/TE* = 2000*ms/*30*ms*, 90 flip angle, 38 slices with 3 *mm* thickness, field of view (FOV) of 240 × 240 *mm*^2^, matrix size of 64 × 64, and voxel size of 3.75 × 3.75 × 3 *mm*^3^. For each subject, the duration of the rs-fMRI scan was 800 seconds (13 minutes, 20 seconds). The subjects were instructed to remain as still as possible during the resting-state scans with their eyes open.

The Perfusion MRI scans were acquired using the 2D EPI Pseudo Continuous Arterial Spin Labelling (PCASL) sequence with online contrast generation between the control and tagged volumes. The FOV was set to 240 × 240 *mm*^2^ with 42 slices of 4 *mm* thickness. The voxel size was 1.88 × 1.88 × 4 *mm*^3^, and *TR/TE* = 4.728*s/*10.70*ms*. An inversion time of 2.975 seconds was used with a labeling duration of 1.45 seconds and a post-labeling delay of 1.525 seconds.

### 2.4. Analysis of Brain Morphology

T1-weighted MRIs were utilized to study the morphological characteristics of brain anatomy, such as volume and tissue thickness. After a quality check of the images, we included the T1-weighted MRI of 75 CRPs (32.12 *±* 11.37 years, 18 Female) and 50 HCs (31.68 *±* 8.79 years, 12 Female) in this analysis.

#### 2.4.1. Preprocessing

The T1-weighted anatomical MRIs were processed using Freesurfer [15] to remove the skull and non-brain tissue (skull-stripping). Further, to derive morphological features from the structural MRIs, we used a combination of Freesurfer and FSL [16] algorithms.

#### 2.4.2. Whole-Brain Morphological Features

We used FMRIB’s Automated Segmentation Tool (FAST) to extract whole-brain morphological features like total intracranial volume (TIV), grey matter volume (GMV), white matter volume (WMV), and cerebrospinal fluid volume (CSFV) [17]. We also defined the total brain tissue volume (BTV) as the TIV without including the volume of ventricles. In addition, the GMV, WMV, and CSFV were also normalised by the TIV to eliminate the confound of head size. All the features were quantile normalised to aid comparison between groups.

#### 2.4.3. ROI-Based Morphological Features

Cortical surface reconstruction was performed using Freesurfer 6.0.0 to get the morphological features at the level of Regions of Interest (ROIs) in the cortex. These features include cortical volume and cortical thickness of 68 ROIs defined using the Desikan-Killiany atlas [18] along with the mean of each hemisphere. Volume of subcortical ROIs in the brain was also extracted using the FMRIB’s Integrated Registration and Segmentation Tool (FIRST) algorithm in the FSL Toolbox. The volumes of all cortical and subcortical ROIs were normalised by the whole-brain volume (TIV) for comparison across subjects. All the cortical and sub-cortical features were quantile normalised to aid comparison between groups.

#### 2.4.4. Surface-Based Morphometry (SBM)

Cortical reconstruction using FreeSurfer 6.0.0, the “recon-all” processing pipeline was utilized to extract surface-based morphological features from T1-weighted MRI scans [19, 20]. The individually reconstructed surfaces were inflated and mapped to a common spherical coordinate system, “fsaverage”, facilitating precise inter-subject alignment. A map of cortical thickness (CT) was obtained by calculating the distance between the pial and white matter surfaces at each vertex [21]. Additionally, a cortical volume (CV) map was derived by integrating cortical thickness and surface area measurements, quantifying regional variations in GM architecture [22]. These CT and CV maps were derived for all subjects and compared to study the granular changes in brain morphology.

#### 2.4.5. Statistical Analysis

We used non-parametric tests for statistical comparison of feature distributions due to potential violations of normality assumptions. Specifically, the two-sided Mann–Whitney U test was used as a non-parametric alternative to the two-sample t-test for group comparisons. Features within each domain were tested together and corrected for multiple comparisons using the Benjamini–Hochberg False Discovery Rate (FDR-BH) method, with statistical significance set at *p*_FDR_ < 0.05. Whole-brain morphological features, including TIV, BTV, GMV, WMV, CSFV, and their TIV-normalized variants, were assessed collectively. Volumes of subcortical ROIs (*N* = 15), cortical ROIs (*N* = 70), and cortical thickness values (*N* = 70) were similarly compared using Mann–Whitney U tests and corrected using FDR-BH.

For SBM, a generalized linear model (GLM) was applied to compare CT and CV maps between HCs and CRPs, incorporating age and sex as covariates of no interest. Cluster-level statistical significance was determined using FDR correction at *p*_FDR_ < 0.05 (*p*_*unc*_ < 0.001) to account for multiple comparisons across surface vertices.

#### 2.4.6. Relationship with Infection Severity

T1-weighted MRIs of 50 HCs (31.68 *±* 8.79 years, 12 Female), 20 HPs (38.85 *±* 11.90 years, 5 Female), and 46 NHPs (27.30 *±* 7.72 years, 10 Female) passed the quality checks and were included in this analysis. To investigate the effect of infection severity on whole-brain and ROI-based morphological measures, we performed non-parametric Kruskal-Wallis H-test for each set of measures. The features were grouped identically as described for the cohort comparison. The p-values were corrected for FDR using the BH method at *p*_*F DR*_ < 0.05. Further, for measures that showed a significant effect of severity, post-hoc tests were performed. Dunn’s test was utilised for the post-hoc pairwise comparison of groups with multiple comparison corrections for FDR using the BH method at *p*_*F DR*_ < 0.05.

Similar to the comparison of HCs and CRPs, a GLM was used to study the effect of severity on the CT and CV maps with a covariate each for HCs, NHPs, and HPs. Again, age and sex were added as covariates of no interest. Significant clusters were identified using an FDR-corrected threshold of *p*_*F DR*_ < 0.05 (*p*_*unc*_ < 0.001).

### 2.5. Analysis of White Matter Microstructure

Microstructural properties of the white matter tissue were studied using dMRI. However, after a quality check, we excluded four CRPs and one HC due to poor image quality and finally included 72 CRPs (32.17± 11.51 years, 17F) and 50 HCs (31.68 ± 8.79 years, 12F) in this analysis. The diffusion tensor model was used to derive measures of microstructural integrity like Fractional Anisotropy (FA), Mean Diffusivity (MD), Axial Diffusivity (AD), and Radial Diffusivity (RD).

#### 2.5.1. Preprocessing

The MRtrix3 software was employed to denoise the dMRI scans, followed by the removal of phase-encoding-induced warping, and Gibbs’ ringing artefacts [23]. The susceptibility-induced distortion artefacts were removed using a combination of the Synb0-DISCO algorithm (to synthesise images with opposite phase-encoding) [24] and FSL’s TOPUP algorithm [25]. Further, head motion and Eddy current correction were performed using FSL. VistaSoft software (https://github.com/vistalab/vistasoft) for MATLAB was employed for diffusion tensor reconstruction using the ‘dtiInit’ function. dMRI scans were registered to the ACPC-aligned native T1 space along with a corresponding rotation of gradient vectors. The diffusion tensors were estimated using a least-squares algorithm, bootstrapped 500 times.

#### 2.5.2. Whole-Brain Tractography

For tracing the white matter pathways in dMRI, we performed whole-brain deterministic tractography using the Automated Fiber Quantification (AFQ) algorithm in MATLAB [26]. The algorithm was seeded in the white matter and used a fourth-order Runge-Kutta integration method with a 1 mm step size for tracking. The stopping criteria were set to (i) FA < 0.2, or (ii) turning angle > 30. After tractography, the algorithm also performed tract segmentation and fibre tract identification using a probabilistic atlas. Overall, 20 white matter tracts were identified in each subject. To quantify white matter microstructural integrity, diffusion metrics FA, AD, MD, and RD were determined for each tract.

The 20 tracts included the left and right Anterior Thalamic Radiation (ATR), Corticospinal Tract (CST), Cingulum Cingulate (CC), Cingulum Hippocampus (CH), Inferior Fronto-Occipital Fasciculus (IFOF), Inferior Longitudinal Fasciculus (ILF), Superior Longitudinal Fasciculus (SLF), Uncinate Fasciculus (UF), Arcuate fasciculus (AF), along with major and minor Callosum Forceps (CF).

#### 2.5.3. Statistical Analysis

The diffusion measures (FA, MD, AD, RD) of 20 white-matter tracts were compared across the two cohorts without the assumption of normality using the Mann-Whitney U test (N = 80). FDR was controlled using the FDR-BH method at *p*_*F DR*_ < 0.05.

#### 2.5.4. Relationship with Infection Severity

dMRI scans of 50 HCs (31.68 ± 8.29 years, 12F), 20 HPs (38.85 ± 11.90 years, 5F), and 44 NHPs (27.36 ± 7.86 years, 9F) passed the quality check and were included in this analysis. To study the effect of infection severity on the diffusion measures (FA, MD, AD, RD) of 20 tracts, we conducted Kruskal-Wallis H-tests. Multiple-comparison correction was performed using the FDR-BH method at *p*_*F DR*_ < 0.05. For tracts that showed a significant effect of severity, post-hoc comparisons between groups relied on Dunn’s Test with multiple comparison corrections for FDR using the BH method at *p*_*F DR*_ < 0.05.

### 2.6. Analysis of Brain Function

We studied functional connectivity using the resting state fMRI scans. From the total set of subjects, we excluded 16 CRPs and 4 HCs due to poor-quality images and motion artifacts, leading to a sample size of 61 CRPs (32.02 ± 11.01 years, 14F) and 47 HCs (32.30 ± 9.23 years, 11F) for this study. We utilised Independent Components Analysis (ICA) to extract maps of functionally connected regions in the brain.

#### 2.6.1. Preprocessing

The fMRI data were preprocessed using the Statistical Parametric Mapping (SPM) toolbox in MATLAB (The MathWorks, Inc., Natick, MA, USA). The volumes were corrected for head motion using 6-parameter rigid body transformations to align them to the mean functional image. Subjects with a frame-wise displacement of greater than 2 *mm* in any slice of the run were excluded from the study. The mean image of the motion-corrected functional volumes was co-registered to the anatomical (T1-w) image. The anatomical images were segmented to extract grey-matter (GM), white-matter (WM), and cerebrospinal-fluid (CSF) tissue probability maps (TPMs) along with a population average anatomical template using DARTEL [27]. The TPMs were non-linearly warped to the MNI standard template. Subsequently, the co-registered functional volumes were normalized to the MNI space. Spatial smoothing was done using a Gaussian kernel with *FWHM* = 8 *mm*. The WM and CSF nuisance signals were extracted. The nuisance signals, along with the six motion parameters, were regressed out from the fMRI time series. Temporal filtering was performed using a Butterworth band-pass filter with cut-offs at 0.01 Hz and 0.1 Hz. The functional images were then resampled to an isotropic resolution of 3mm.

#### 2.6.2. Independent Component Analysis of rs-fMRI

The BOLD time-series of all subjects were concatenated in time and flattened into a 2D matrix. Group Independent Component Analysis (ICA) of this matrix was used to extract the predominant resting-state networks (RSNs) across all subjects. FSL’s MELODIC Tool [28] was used to implement this algorithm, extracting 20 independent components (ICs). Using AFNI’s 3dMatch tool, the ICs were classified into canonical RSNs based on their Dice coefficient similarity with the respective networks in the Yeo Atlas (17-Networks) [29]. Among the 20 ICs, 14 ICs with dice coefficients greater than 0.254 were identified as canonical RSNs, which are shown in Figure 1. Dual Regression [30] was performed to obtain subject-wise time courses and spatial maps for each identified IC. The subject-specific maps of RSNs were compared across the cohorts to check for group-level differences.

**Figure 1:**
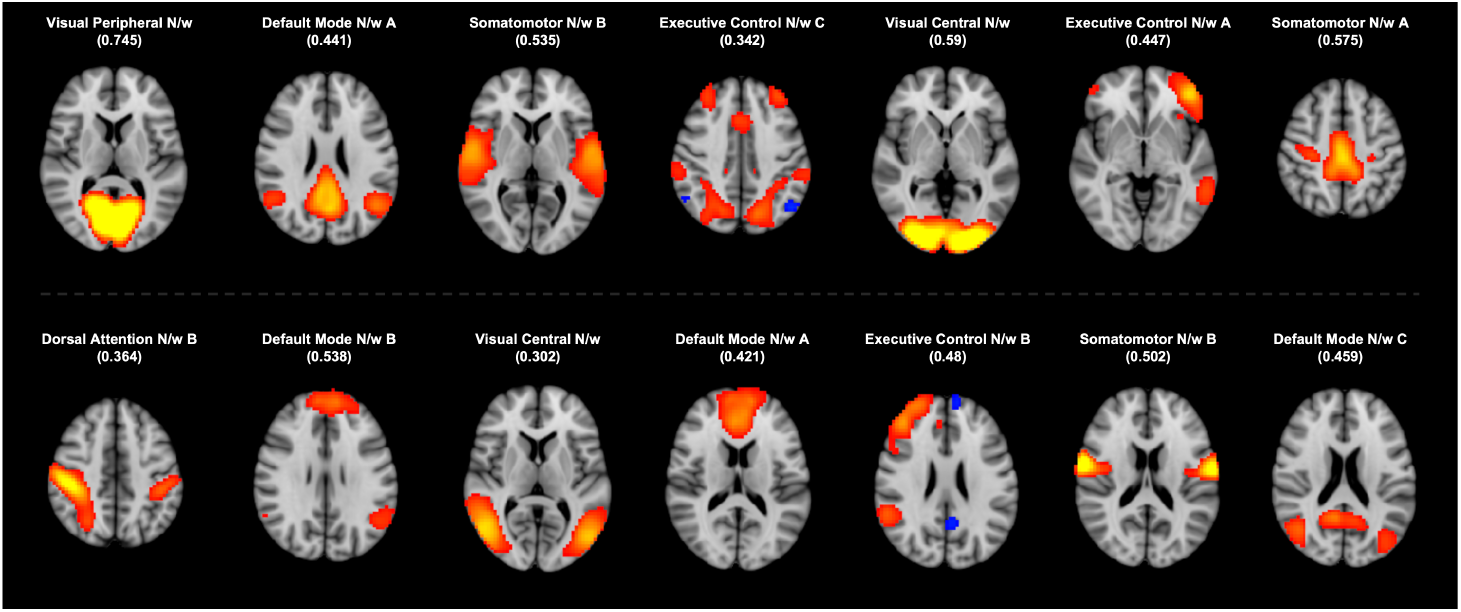
Fourteen Resting State Networks (RSN) were identified by comparing the independent components (ICs) with network maps from the Yeo Atlas (https://surfer.nmr.mgh.harvard.edu/fswiki/CorticalParcellation_Yeo2011 ). The identified ICs are presented here with the best-matched RSN from the atlas. The values of corresponding correlation coefficients are provided in parentheses. Key: N/w = Network

#### 2.6.3. Statistical Analysis

For each IC that was identified as a canonical RSN, we compared the subject-wise spatial maps of the network across the two cohorts using a permutation test. FSL’s randomise was used to conduct the test with 5000 permutations. Correction for multiple comparisons was done using the Threshold Free Cluster Enhancement (TFCE) method at *p <* 0.05 [31].

#### 2.6.4. Relationship with Infection Severity

The effect of infection severity on resting-state functional connectivity was studied using 47 HCs (32.30 ± 9.23 years, 11F), 14 HPs (41.21 ± 12.59 years, 5F), and 39 NHPs (27.49 ± 8.24 years, 7F). For each IC that was identified as a canonical RSN, we compared the subject-wise spatial maps of the three groups using permutation testing (N = 5000). Correction for multiple comparisons was performed using the TFCE method at *p <* 0.05.

### 2.7. Analysis of Cerebral Blood Flow

Blood perfusion in the brain can be detected by using the Arterial Spin Labelling (ASL) MRI technique. Here, a contrast is generated in the blood vessels of the brain by magnetically tagging incoming blood with an inverted spin [32]. For the analysis of cerebral blood flow (CBF) using ASL images, we included 67 CRPs (32.21 ± 11.82 years, 15F) and 47 HCs (31.98 ± 8.95 years, 11F), excluding 9 CRPs and 4 HCs due to poor image quality.

#### 2.7.1. Data Processing

With the ASL protocol described in earlier sections, difference images (tagged–control) were obtained. The data was processed using the Oxford ASL pipeline in FSL toolbox [33]. Preprocessing involved motion correction and spatial regularisation. Anatomical images processed using the FSL-ANAT pipeline [16] were utilised to co-register the pre-processed difference images to native space, followed by normalisation to MNI space using FSL. The obtained CBF values were calibrated using the M0 image to get the CBF in absolute units. Partial volume correction was then done to obtain separate perfusion maps for grey and white matter. These perfusion maps were used to perform a region analysis based on the ROIs defined in the Harvard-Oxford Atlas. Therefore, perfusion values for 68 ROIs were obtained and used for statistical comparison.

#### 2.7.2. Statistical Analysis

Perfusion values were obtained for 68 ROIs. For each ROI, these values were compared across CRPs and HCs using the Mann-Whitney U Test. The results were corrected for multiple comparisons using the FDR-BH method at *p*_*F DR*_ < 0.05.

#### 2.7.3. Relationship with Infection Severity

CRPs were classified into NHPs and HPs to study the effect of severity on cerebral blood flow. We included 47 HCs (31.98 ± 8.95 years, 11F), 42 NHPs (27.12 ± 7.84 years, 8F), and 17 HPs (40.23 ± 12.21 years, 4F) in this analysis. The comparison was done using the Kruskal-Wallis H-test. Multiple-comparison correction was performed using the FDR-BH method at *p*_*F DR*_ < 0.05.

## 3. Results

### 3.1. Subject Demographics and Characteristics

For this study, we recruited a total of 127 subjects. The demographic details of the cohorts included in our analysis are presented in Table 1. In the CRP group, 67 subjects consented to sharing their treatment history. Based on treatment history 46 CRPs were classified as NHPs. The other 21 patients were classified as HPs and had undergone hospitalisation during their treatment, requiring oxygenation (10/21), administered remdesivir (10/21), or steroids (3/21). The most commonly experienced symptoms for the CRPs were fever (85.07%) and cough (70.15%), followed by body ache (64.17%), loss of smell (44.77%), chills (37.31%), loss of taste (37.31%), breathing difficulties (34.33%), nausea (19.40%), and weakness (14.92%). Figure 2 (a) presents a summary of the range of symptoms that were experienced by the CRPs in our study.

**Table 1:**
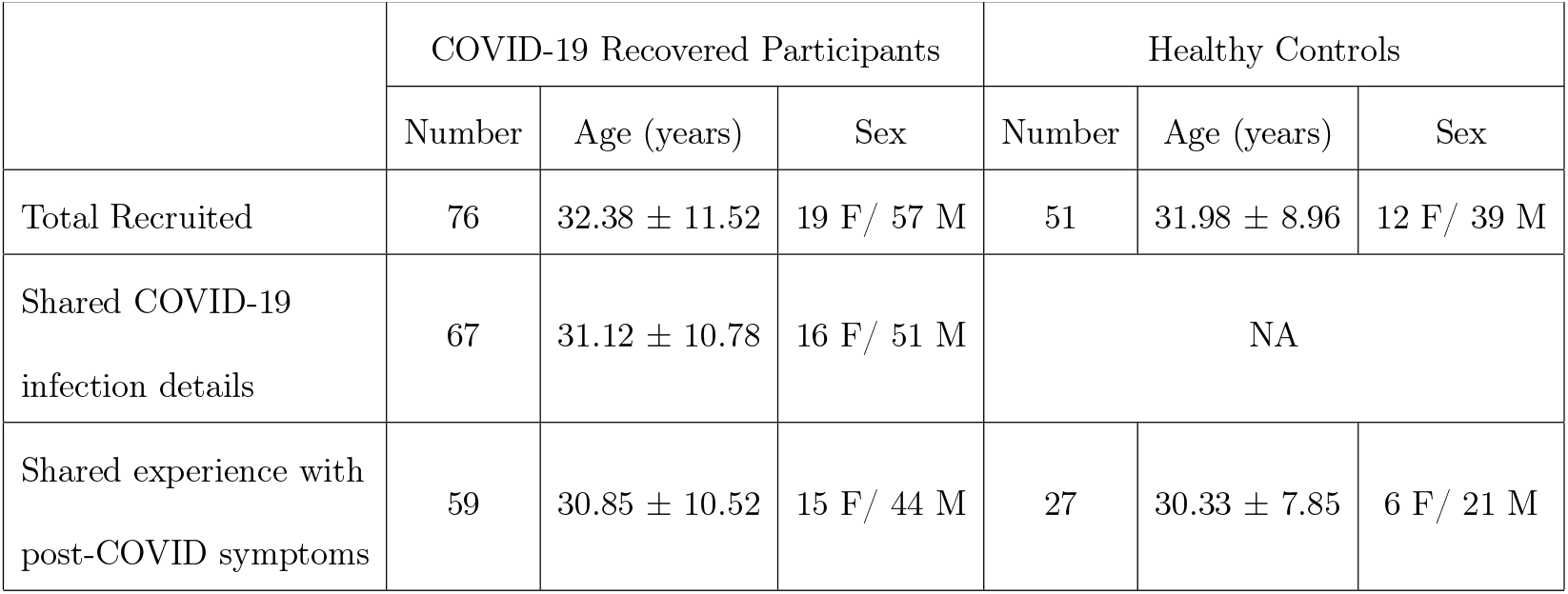
Demographic details of the participants recruited for this study. The age distribution is represented using the “mean ± standard deviation” values.

**Figure 2:**
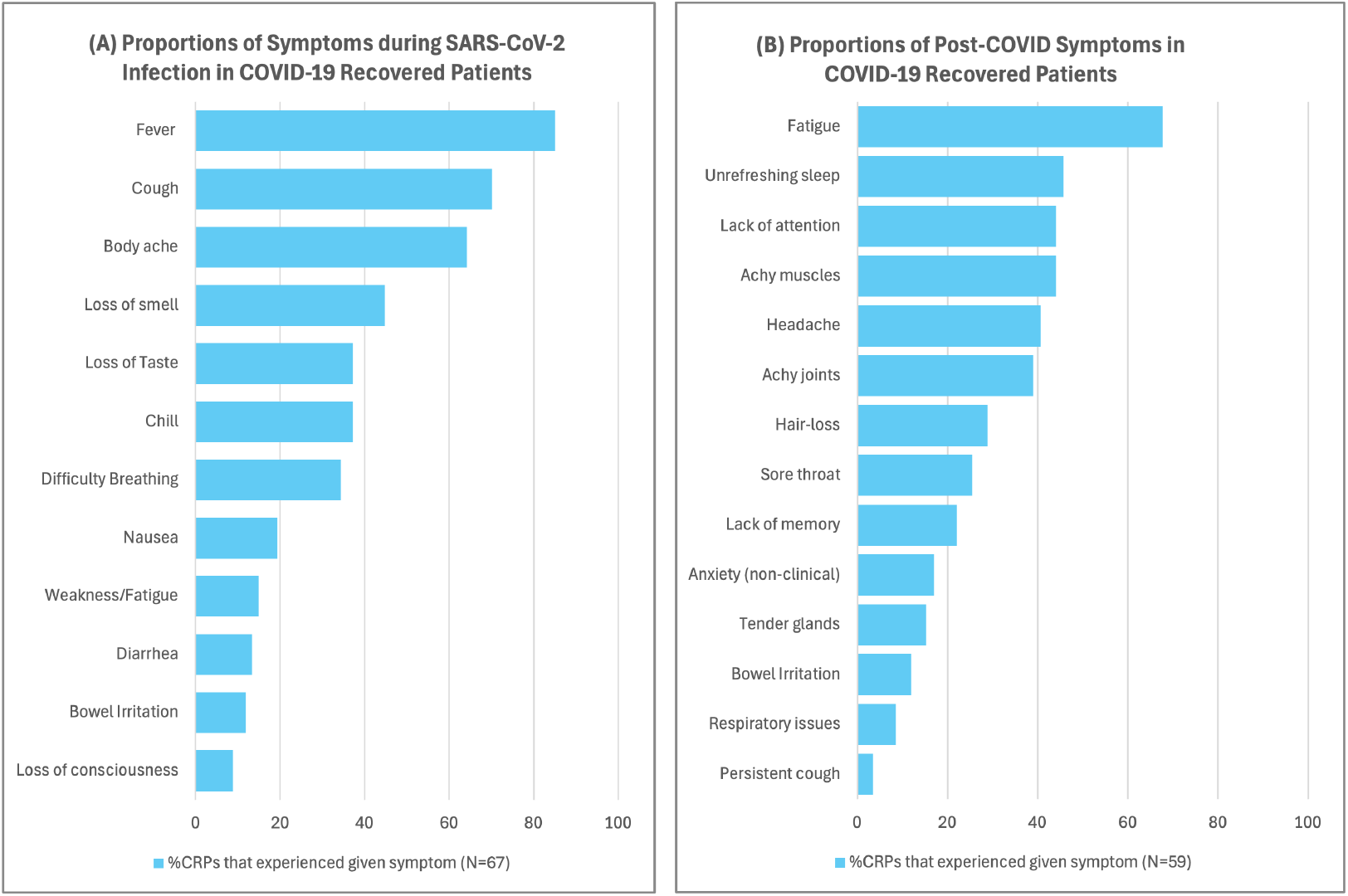
(a) Proportions of Symptoms during SARS-CoV-2 Infection in COVID-19 Recovered Patients, total CRPs = 67. (b) Proportions of Post-COVID Symptoms in COVID-19 Recovered Patients.

We also requested CRPs to share their post-COVID experience and any persistent symptoms since recovery. Among 59 (30.85 ± 10.52 years, 15F) consenting CRPs, fatigue (40/59) was the most commonly reported post-COVID symptom, followed by unrefreshing sleep (27/59) and lack of attention (26/59). Other symptoms reported by CRPs were achy muscles (26/59), achy joints (23/59), and headache (24/59). A summary of the post-COVID symptoms is presented in Figure 2 (b).

### 3.2. Differences between COVID-19 Survivors and Controls

#### 3.2.1. No Difference in Whole-Brain and Cortical Morphology Between Cohorts

First, as a baseline, we performed a comparison of the whole-brain morphological features. We found no significant differences in TIV, BTV, GMV, WMV, CSFV, GMVnorm, WMVnorm, or CSFVnorm between CRPs and HCs (*p*_*F DR*_ < 0.05). Further, we compared the morphological features of the cortical ROIs among the cohorts. The cortex was segmented into 68 ROIs based on the Desikan-Killiany atlas, and the volume and average thickness of the 68 ROIs, along with the mean thickness and volume of each hemisphere, were determined. Comparison of ROI volumes between the cohorts yielded no significant differences (*p*_*F DR*_ < 0.05). Moreover, the thickness of the cortical ROIs also showed no significant differences when compared across the CRP and HC cohorts (*p*_*F DR*_ < 0.05).

#### 3.2.2. Reduced ROI Volume in Right Basal Ganglia and Limbic System

The volumes of 15 subcortical ROIs were computed for each subject and normalised to the TIV. Statistical comparison of the subcortical ROI volumes indicated significant differences in six regions (*p*_*F DR*_ < 0.05) in the right hemisphere. The regions identified were the thalamus, caudate, putamen, pallidum, hippocampus, and the amygdala. All the ROIs showed reduced volume in CRPs as compared to HCs. It was observed that the putamen, caudate nucleus, and pallidum are essential regions of the basal ganglia in the brain. Moreover, the thalamus, hippocampus, and amygdala are crucial components of the limbic system. Figure 3 shows the distribution of subcortical volumes for the ROIs that exhibited a significant difference between the cohorts.

**Figure 3:**
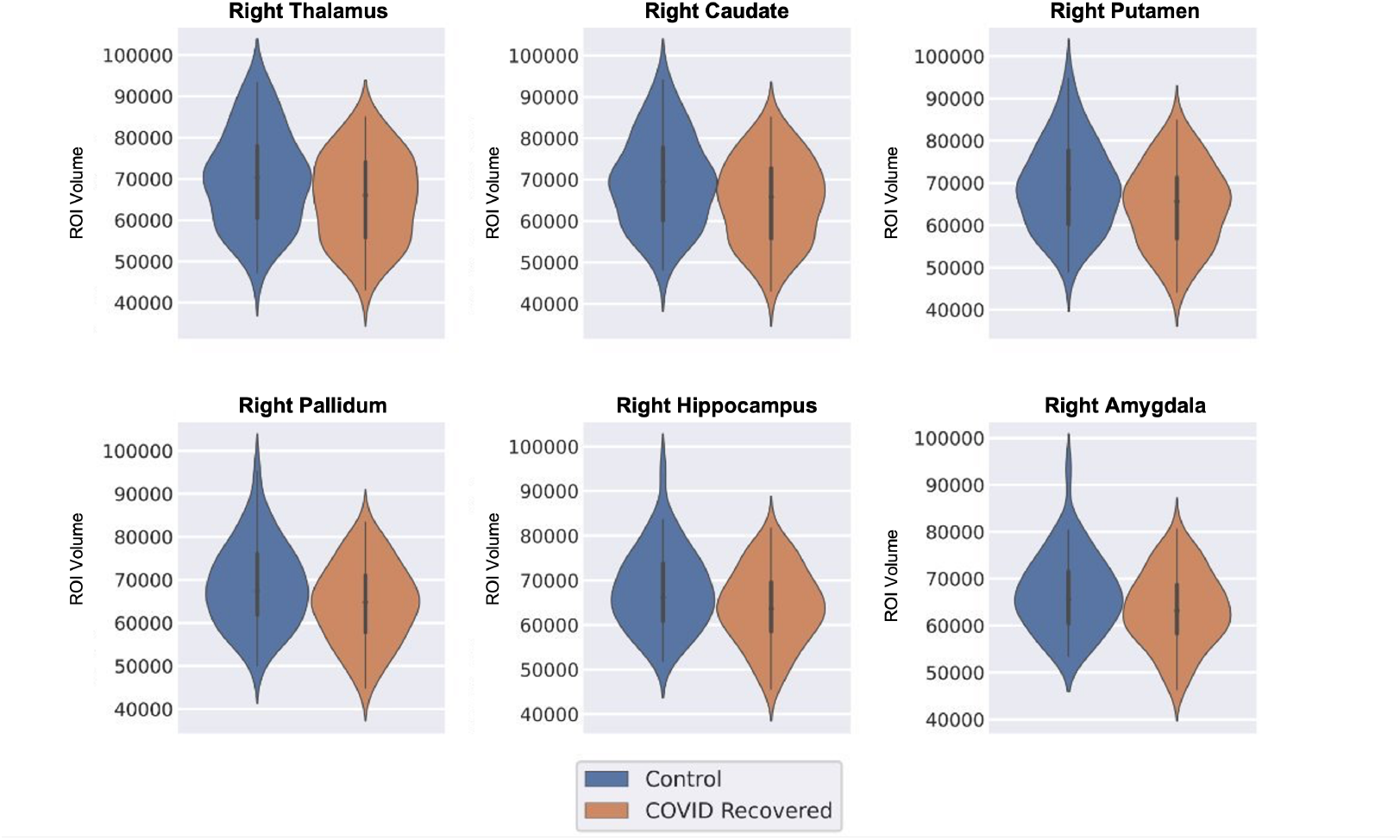
Distribution of volume of regions that showed significant differences between the COVID-19 Recovered Patients (CRPs) and Healthy Controls (HCs) (*p*_*F DR*_ < 0.05)

#### 3.2.3. CRPs Show Reduced Grey-Matter Volume

Using SBM, the thickness and volume of grey matter tissue in the cortex were compared across HCs and CRPs over a reconstructed surface. A GLM was used to test the changes at each vertex of the surface. *p*-values were corrected for multiple comparisons using the FDR method.

Comparison of cortical GM thickness between HCs and CRPs did not highlight any significant clusters (*p*_*F DR*_ < 0.05). On the other hand, comparison of cortical GM volume across the cohorts revealed a significant cluster with reduced volume in CRPs (*p*_*F DR*_ < 0.05, HC>CRP). The cluster over-laps the posterior parts of the right superior temporal gyrus and covered parts of the insular cortex (Figure 4 (A)). Table 2 summarizes the details of the cluster reported here.

**Table 2:** Summary of clusters observed showing significant differences in cortical thickness and cortical volume among the Healthy Controls (HCs), COVID-19 Recovered Patients (CRPs), Non-Hospitalized Patients (NHPs), and Hospitalized Patients (HPs).

| Cortical Thickness |  |  |  |  |  |  |
| --- | --- | --- | --- | --- | --- | --- |
| Contrast | Peak t-score | Size ($mm^2$) | Peak X | Peak Y | Peak Z | Region Identified |
| HC > HP | 3.4556 | 204 | -7.2 | 22.9 | 43.8 | Left Medial Prefrontal Cortex |
| Cortical Volume |  |  |  |  |  |  |
| Contrast | Peak t-score | Size ($mm^2$) | Peak X | Peak Y | Peak Z | Region Identified |
| HC > CRP | 4.222 | 216.62 | 48.7 | -24.1 | 4.8 | Right Superior Temporal Gyrus, Insular Cortex |
| HC > HP | 3.7983 | 197.44 | -6.7 | 24.7 | 42.9 | Left Medial Prefrontal Cortex |

**Figure 4:**
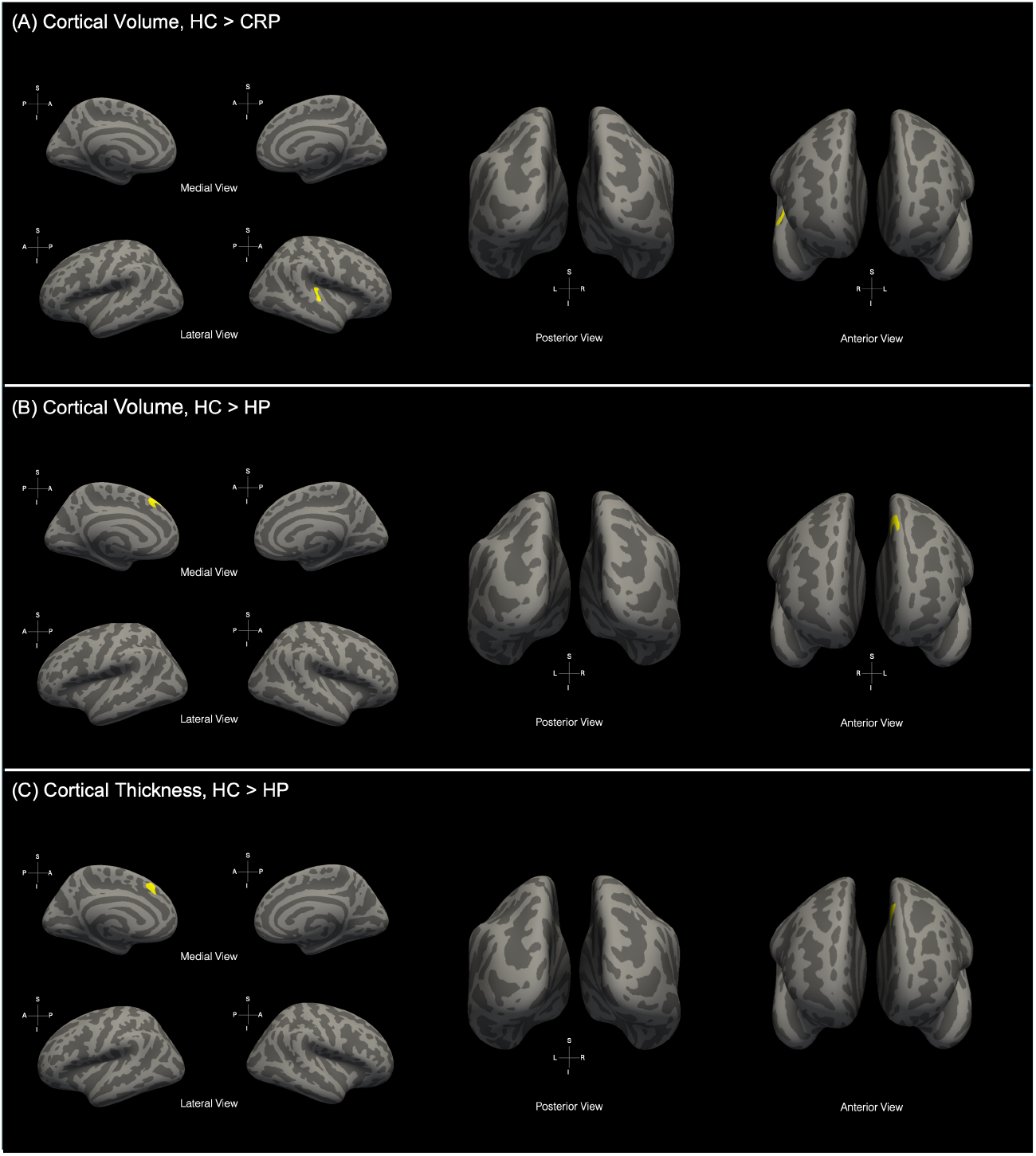
Significant clusters obtained upon comparison of surface-based morphological metrics: (A) Cortical volume differences beween Healthy Controls (HCs) and COVID-19 Recovered Patients (CRPs) where cluster was observed in Right Superior Temporal Gyrus, insular cortex (*p*_*F DR*_ < 0.05, HC>CRP). Significant difference in (B) cortical volume and (C) cortical thickness were observed between HCs and Hospitalized Patients (HP) wherein the cluster was found Left Medial Prefrontal Cortex (*p*_*F DR*_ < 0.05, HC>HP).

#### 3.2.4. Microstructural Differences in Tracts of the Limbic System

The AFQ algorithm was employed to trace and identify 20 white matter tracts in the brain for each subject. For each tract, four measures (FA, MD, AD, RD) were calculated that quantify the microstructural integrity of the tracts. These values were compared between the CRPs and HCs to test for any post-COVID differences in white matter integrity.

We found four tracts that exhibited significant alterations in FA among the groups (*p*_*F DR*_ < 0.05). These results are consistent with the observations on the subset of this data, as reported in [34]. The left UF and the right CC showed reduced values of FA in the CRPs compared to HCs. On the other hand, the right CH and the right ILF had greater FA values in CRPs. Further, significant aberrations in MD were observed in three white matter tracts, including the left CC, left UF, and right CH (*p*_*F DR*_ < 0.05). The left UF exhibited higher MD in the CRPs as compared to HCs, while MD was lower in the CRPs for left CC and right CH. The comparison also highlighted significant alterations in RD for left UF, right CH, and right ATR (*p*_*F DR*_ < 0.05). RD was elevated in the CRPs as compared to HCs for the left UF and right ATR, while it showed a decrease for the right CH. Furthermore, two tracts also showed significant differences in AD values between the two cohorts (*p*_*F DR*_ < 0.05). We found that the left and right CC had reduced AD in the CRPs as compared to the HCs. Figure 5 shows the violin plots of the distribution of diffusion measures described above.

**Figure 5:**
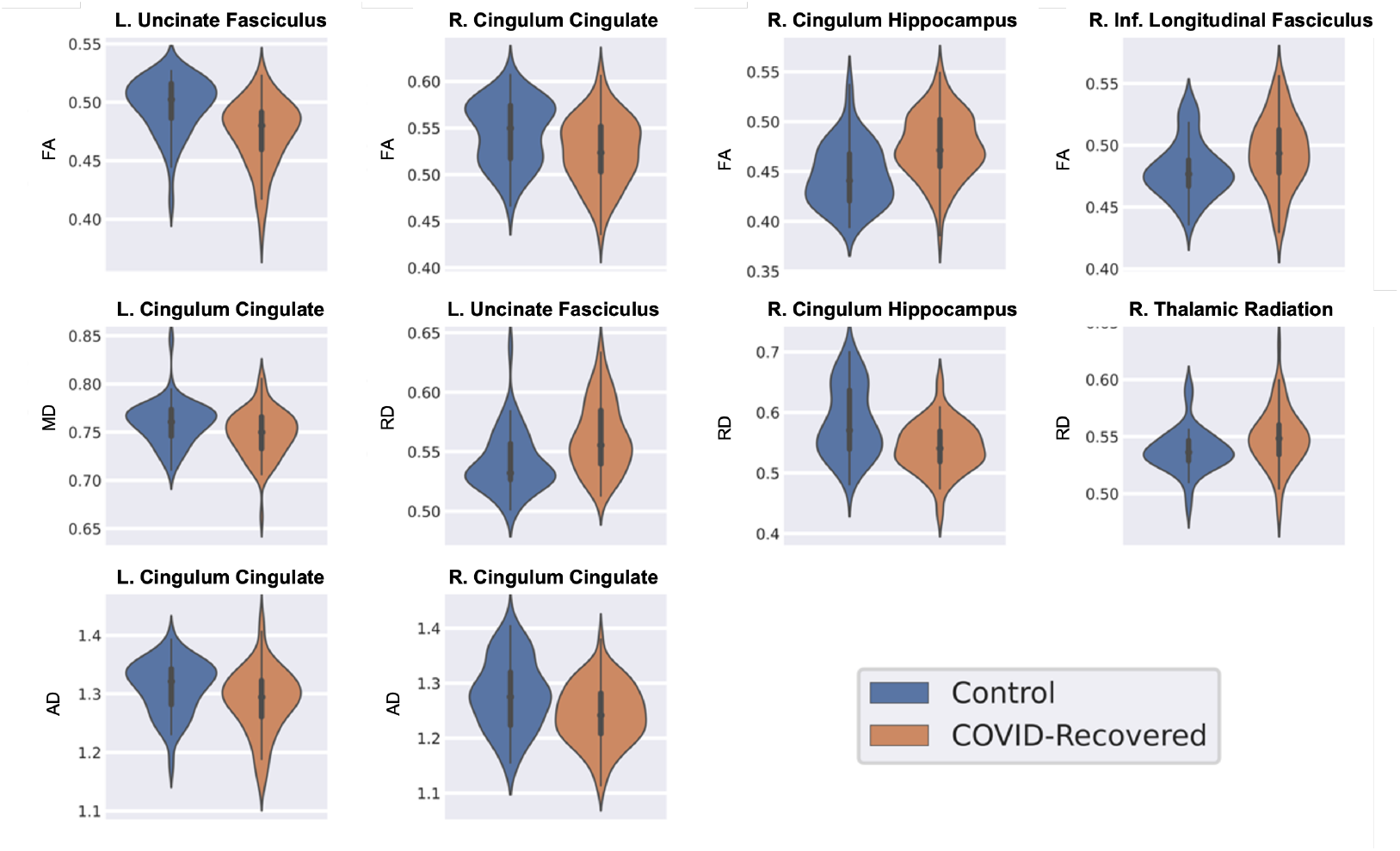
Distribution of diffusion metrics of tracts that showed a significant difference between Healthy Controls (HCs) and COVID-19 Recovered Patients (CRPs) (*p*_*F DR*_ < 0.05). Key: L. = Left, R. = Right, FA = Fractional Anisotropy, MD = Mean Diffusivity, AD = Axial Diffusivity, RD = Radial Diffusivity.

#### 3.2.5. No Group Differences in Activity of Resting-State Networks

FSL’s MELODIC tool was used to extract 20 ICs across all subjects. Among these, 14 ICs were identified as canonical RSNs using Dice coefficients. ICs with high spatial overlap with RSNs from the Yeo Atlas were labelled and included in the statistical comparison. The identified ICs are in Figure 1 along with respective dice coefficients. For each identified IC, we compared the subject-wise spatial maps across the two cohorts using a permutation test. The test revealed no significant cluster with p<0.05 in any of the RSNs.

#### 3.2.6. No Significant Group Differences in Cerebral Blood Flow

We compared the average blood perfusion values for 68 ROIs across HCs and CRPs. We did not find significant differences in the perfusion values in any of the ROIs at *p*_*F DR*_ < 0.05. The ROI that showed the strongest contrast between the cohorts was the left pallidum with *p*_*unc*_ = 0.0926.

### 3.3. Effect of Severity of COVID-19 Infection

Next, we investigated whether these brain changes were associated with the severity of the COVID-19 infection. Based on available treatment history of 67 CRPs, 46 CRPs were classified as NHPs. The other 21 patients who had undergone hospitalisation during their treatment (requiring oxygenation, administered remdesivir, or steroids) were classified as HPs.

#### 3.3.1. Whole-Brain Morphology Not Linked to Severity

The Kruskal-Wallis H-test detected no significant effect of severity within HC, NHP, and HP groups for whole brain features - TIV, BTV, GMV, WMV, CSFV, GMVnorm, WMVnorm, or CSFVnorm (*p*_*F DR*_ < 0.05). As no significant changes linked to severity were observed, post hoc tests were not performed.

#### 3.3.2. ROI-Level Morphological Changes Linked to Infection Severity

The Kruskal-Wallis H-test was used to test the effect of infection severity on the cortical volume and thickness across 70 cortical ROIs. No significant effect of severity was observed in the case of the cortical ROIs defined using the Desikan-Killiany atlas (*p*_*F DR*_ < 0.05).

However, a significant effect of infection severity on the subcortical ROIs (*p*_*F DR*_ < 0.05) was observed. The effect of severity on sub-cortical volume was significant in the right thalamus, right caudate, right putamen, right pallidum, right amygdala, and the right hippocampus (*p*_*F DR*_ < 0.05). Post-hoc Dunn’s tests were conducted for these ROIs. For all six ROIs tested, it was noted that the NHPs had significantly reduced volume as compared to the HCs (*p*_*F DR*_ < 0.05), while no significant differences were observed upon comparing HCs and HPs. Additionally, in the case of the thalamus and caudate, the NHPs had significantly less volume as compared to the HPs. Figure 6 shows the violin plots of the distribution of subcortical volumes for the ROIs that exhibited a significant effect of infection severity.

**Figure 6:**
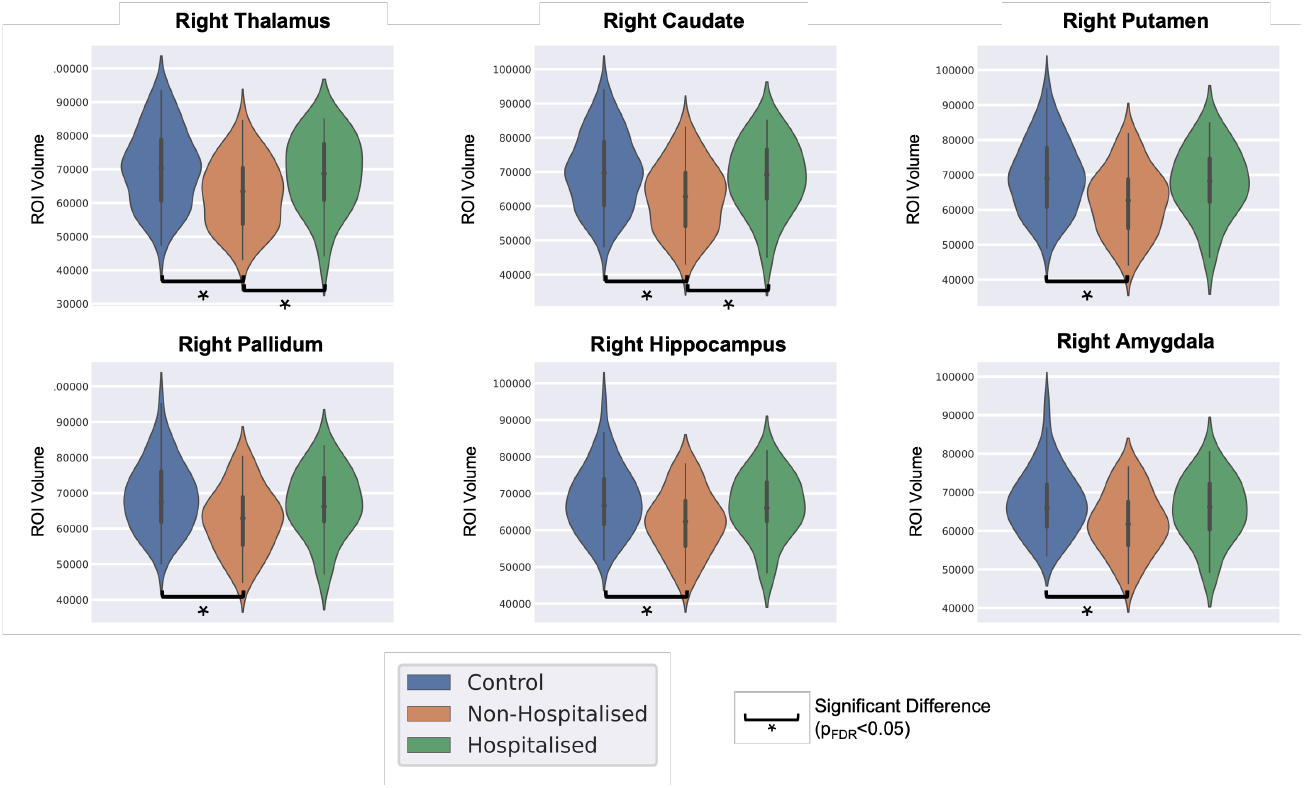
Distribution of volume of regions that showed a significant effect of infection severity among Healthy Controls (HCs), Non-Hospitalized Patients (NHPs), and Hospitalized Patients (HPs) (*p*_*F DR*_ < 0.05).

#### 3.3.3. Grey Matter Thickness and Volume Linked with Infection Severity

The effect of infection severity on GM thickness and volume was tested using a GLM. Using an F-test, we observed a significant effect of severity on cortical volume in HCs, NHPs, and HPs (*p*_*F DR*_ < 0.05, *p*_*unc*_ < 0.05). Post-hoc and pair-wise tests were conducted to identify the groups with significant differences.

Pair-wise tests on the cortical GM thickness highlighted significant differences between the HC and HP groups. Comparison between these groups yielded a significant cluster in the left medial prefrontal cortex (*p*_*F DR*_ < 0.05, *p*_*unc*_ < 0.001), Figure 4 (B)). Post-hoc comparisons of cortical GM volume also highlighted significant differences. Comparison of HCs with HPs revealed a significant cluster in the left medial prefrontal cortex (*p*_*F DR*_ < 0.05, *p*_*unc*_ < 0.001), Figure 4 (C)). The details of the clusters are presented in Table 2.

#### 3.3.4. Microstructural Differences Between HCs, NHPs, and HPs

The Kruskal-Wallis H-test detected significant differences between HC, NHP, and HP for four tracts (*p*_*F DR*_ < 0.05). The left UF and right CH showed a significant effect of severity in FA and RD; the left CC showed significant changes linked to severity in MD and AD, and the right ILF exhibited a significant effect of infection severity on FA. Figure 7 presents the violin plots of the distribution of diffusion measures across the HCs, NHPs, and HPs for the results described above.

**Figure 7:**
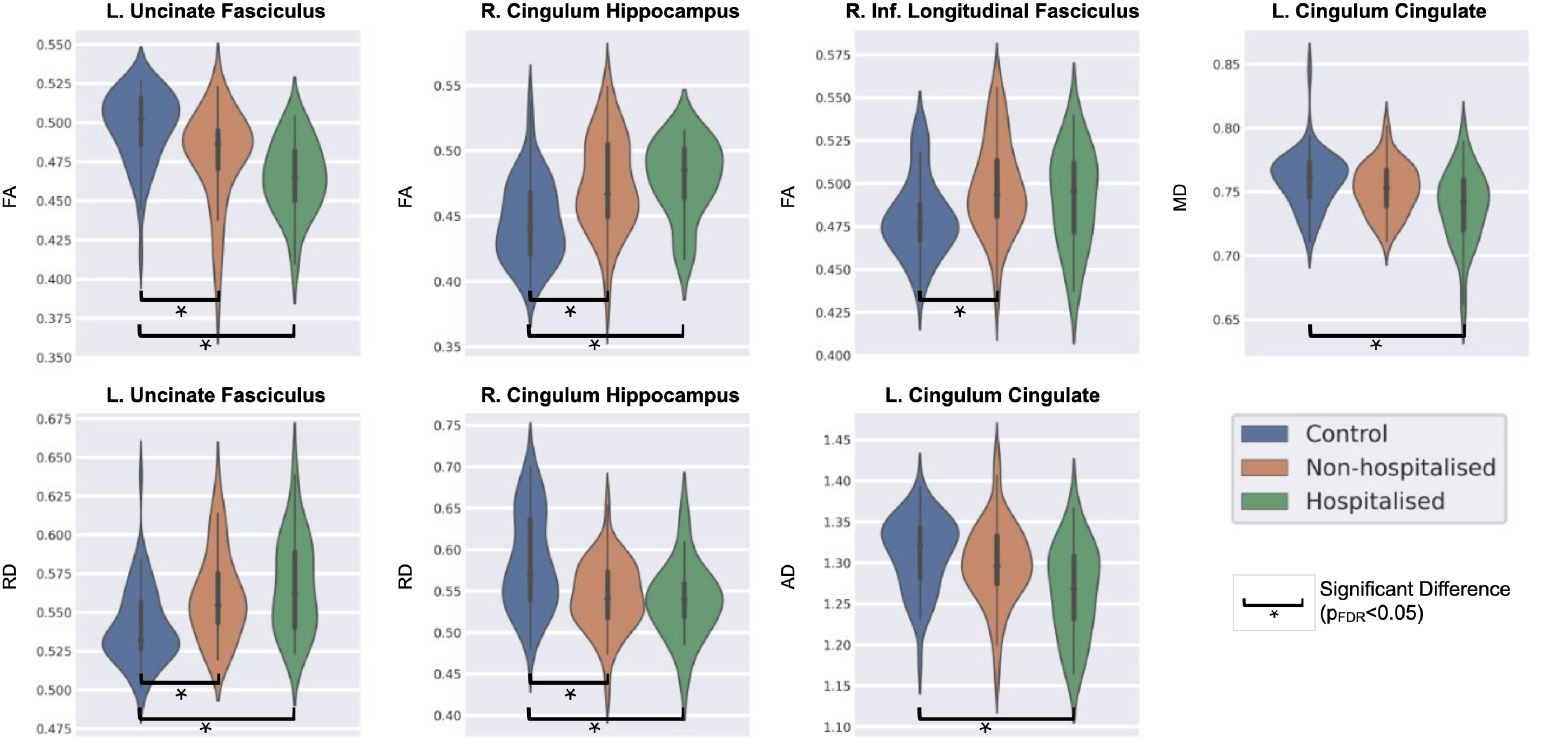
Distribution of diffusion metrics of tracts that showed a significant effect of infection severity amongst Healthy Controls (HCs), Non-Hospitalized Patients (NHPs), and Hospitalized Patients (HPs) (*p*_*F DR*_ < 0.05). Key: L. = Left, R. = Right, FA = Fractional Anisotropy, MD = Mean Diffusivity, AD = Axial Diffusivity, RD = Radial Diffusivity, FDR = False Detection Rate.

Further, upon conducting post-hoc tests on these measures for pairwise comparison between the groups, we found that none of the tracts’ diffusion measures showed a significant difference between the HP and NHP cohorts (*p <* 0.05). Significant pairwise alterations (*p <* 0.05) were, however, observed between the HC and NHP groups in the left UF (FA and RD), right CH (FA and RD), and the right ILF (FA). The left CC did not show significant differences between HC and NHP groups for *p <* 0.05. Additionally, pairwise comparison of HC and HP groups highlighted significant differences (*p <* 0.05) in the left UF (FA and RD), right CH (FA and RD), and the left CC (MD and AD). We did not find any significant differences between the HC and HP groups in the right ILF at a *p <* 0.05 threshold.

#### 3.3.5. Hospitalized Patients Show Significant Functional Changes

Upon testing each of the 14 identified RSNs, we observed a significant effect of infection severity in 4 RSNs (*p*_*corr*_ < 0.05), namely, Default Mode Network – A (DMN-A), DMN-C, Somatomotor Network – A (SMN-A), and the Somatomotor Network - B. Further, we conducted post-hoc pairwise comparisons for these networks between HCs, NHPs, and HPs.

The spatial maps of the DMN-A showed significant differences between the NHPs and HPs (*p*_*corr*_ < 0.05, NHP > HP) in the left posterior insular cortex, which can be observed in Figure 8. Significant clusters were also obtained upon comparing HCs and HPs (*p*_*corr*_ < 0.05, HC > HP) in the same region as reported in Figure 8. Additionally, comparing the HCs and NHPs showed significant differences in the dorsal visual association cortex (*p*_*corr*_ < 0.05, NHP > HC) as shown in Figure 8. Comparison of the spatial maps of the DMN-C between the NHPs and HPs highlighted significant differences in the precuneus, posterior cingulate, and the inferior frontal cortex (*p*_*corr*_ < 0.05, NHP > HP) as shown in Figure 9. Further, significant differences in FC with DMN-C between HCs and HPs were observed in the right caudate and the left thalamus (*p*_*corr*_ < 0.05, HC > HP) as described in Figure 9.

**Figure 8:**
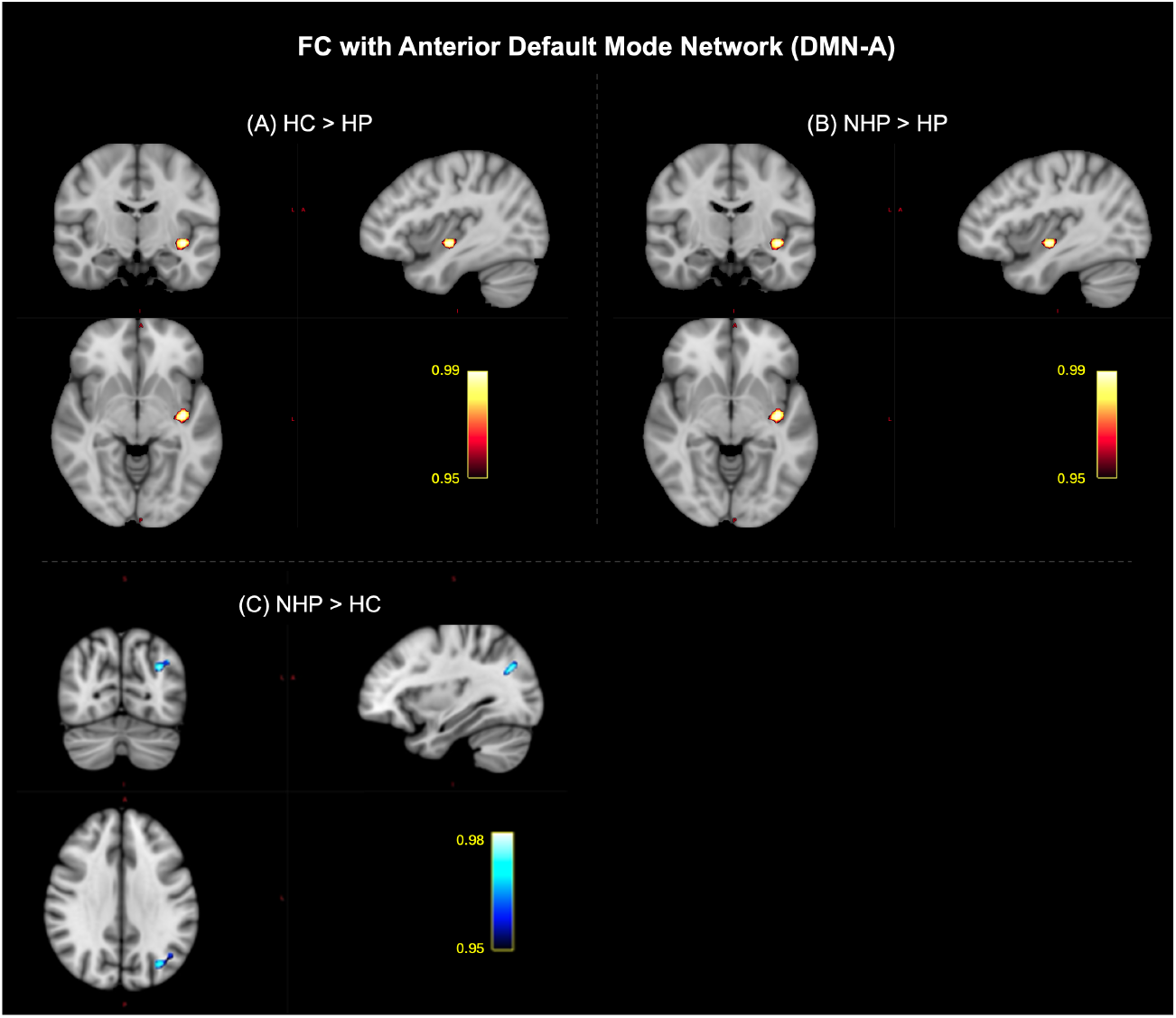
Significant clusters that highlight the effect of infection severity on the functional connectivity (FC) of the anterior default mode network (DMN-A) with the brain amongst Healthy Controls (HCs), Non-Hospitalized Patients (NHPs), and Hospitalized Patients (HPs) (*p*_*corr*_ < 0.05). (A) Post-hoc comparison of HCs and HPs showed a significant cluster in the left posterior insular cortex (HC>HP). (B) Post-hoc comparison of HPs and NHPs also highlighted a cluster in the left posterior insular cortex (NHP>HP). (C) Post-hoc comparison of NHPs and HCs showed a significant cluster in the dorsal visual association cortex (NHP>HC).

**Figure 9:**
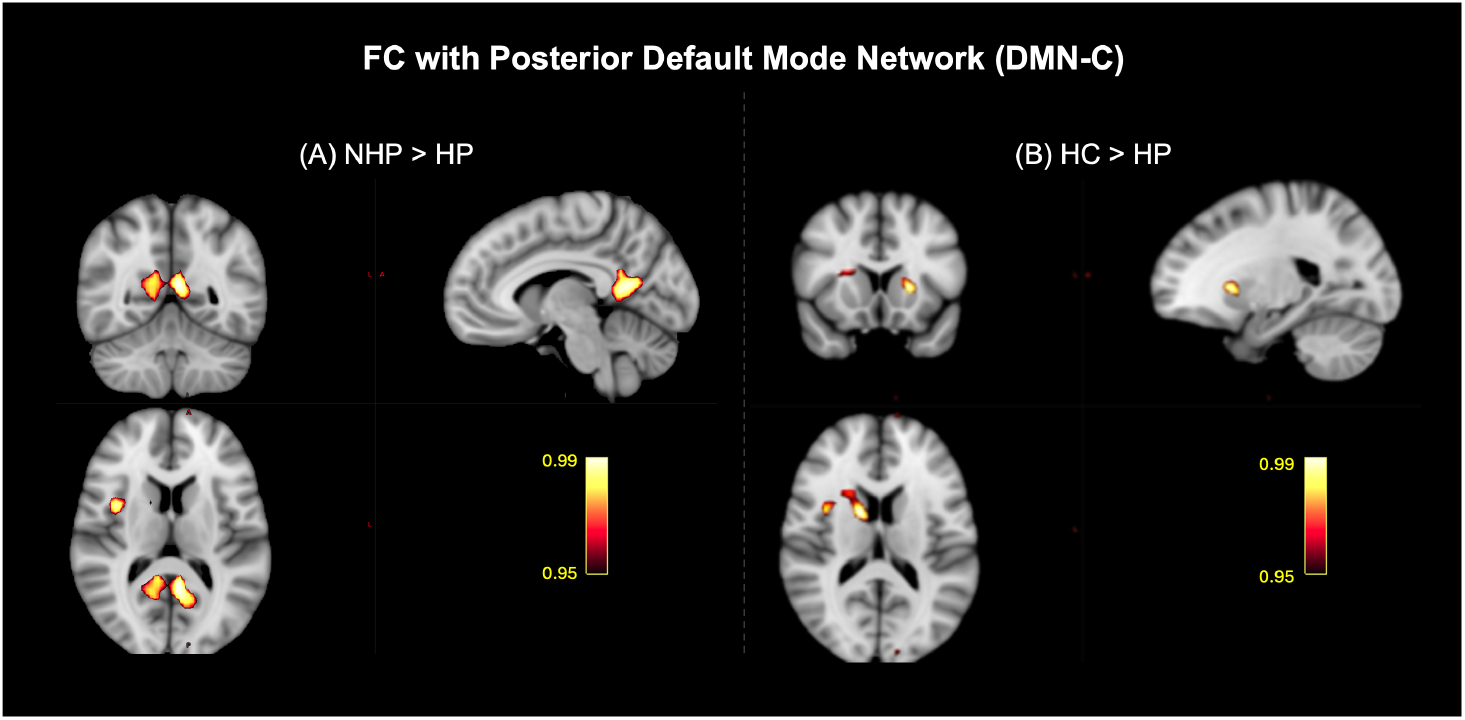
Significant clusters that highlight the effect of infection severity on the functional connectivity (FC) of the posterior default mode network (DMN-C) with the brain amongst Healthy Controls (HCs), Non-Hospitalized Patients (NHPs), and Hospitalized Patients (HPs) (*p*_*corr*_ < 0.05). (A) Post-hoc comparison of NHPs and HPs highlighted clusters in the precuneus, posterior cingulate, and the inferior frontal cortex (NHP>HP), and (B) post-hoc comparison of HCs and HPs showed a significant cluster in the right caudate and the left thalamus (HC>HP).

Spatial maps of the Somatomotor-A network showed significant differences between the NHPs and HPs in the left postcentral gyrus (*p*_*corr*_ < 0.05, NHP > HP, Figure 10). FC with the Somatomotor-B network exhibited significant differences between the NHPs and HPs in the left and right caudate, the right Heschl’s gyrus, and right supramarginal gyrus (*p*_*corr*_ < 0.05, NHP > HP) as shown in Figure 10. For both networks, the comparison of HCs with NHPs and HCs with HPs did not highlight any significant differences (*p*_*corr*_ < 0.05). A summary of the clusters obtained is provided in Table 3.

**Table 3:** Summary of clusters observed showing significant differences in resting-state functional connectivity among the Healthy Controls (HCs), COVID-19 Recovered Patients (CRPs), Non-Hospitalized Patients (NHPs), and Hospitalized Patients (HPs).

| Resting-State Network | Contrast | Peak Confidence (1-p) | Size (Voxels) | Peak X | Peak Y | Peak Z | Region Identified |
| --- | --- | --- | --- | --- | --- | --- | --- |
| Default Mode Network-A | HC >HP | 0.998 | 38 | -36 | -12 | -6 | Left Insular Cortex |
|  | NHP >HP | 0.999 | 100 | -36 | -12 | -9 | Left Insular Cortex |
|  |  | 0.964 | 21 | 3 | -18 | 48 | Cingulate Gyrus, posterior division |
|  | NHP >HC | 0.989 | 49 | -33 | -69 | 30 | Lateral Occipital Cortex, superior division |
| Default Mode Network-C | HC >HP | 0.991 | 87 | 15 | 3 | 15 | Right Caudate, Central Opercular Cortex |
|  |  | 0.991 | 32 | -21 | 15 | 3 | Left Putamen |
|  | NHP >HP | 0.995 | 263 | -6 | -51 | 12 | Precuneus Cortex |
|  |  | 0.992 | 56 | 39 | 3 | 12 | Inferior Frontal Cortex |
| Somatomotor Network-A | NHP >HP | 0.995 | 77 | -45 | -9 | 27 | Postcentral Gyrus |
| Somatomotor Network-B | NHP >HP | 0.994 | 927 | 63 | -42 | 18 | Central Opercular Cortex, Heschl's Gyrus |
|  |  | 0.992 | 164 | -12 | 12 | 9 | Bilateral Caudate |
|  |  | 0.984 | 90 | 54 | -33 | 51 | Supramarginal Gyrus, anterior division |

**Figure 10:**
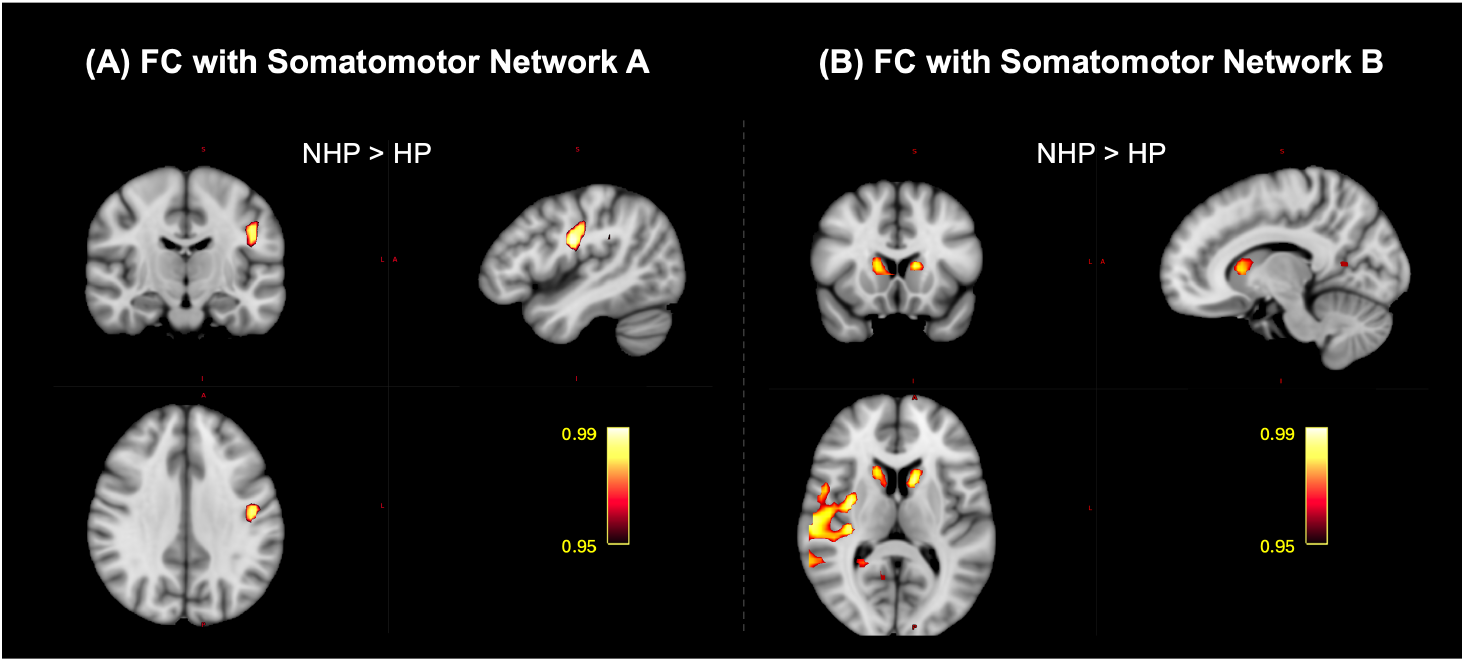
Significant clusters that highlight the effect of infection severity on the functional connectivity (FC) of the somatomotor network (SMN) with the brain amongst Healthy Controls (HCs), Non-Hospitalized Patients (NHPs), and Hospitalized Patients (HPs) (*p*_*corr*_ < 0.05). (A) Post-hoc comparison of the FC of the SMN-A in the NHPs and HPs highlighted a significant cluster in the left postcentral gyrus (NHP>HP), and (B) Post-hoc comparison of the FC of the SMN-B in the NHPs and HPs highlighted a significant cluster in the left and right caudate, the right Heschl’s and supramarginal gyri (NHP>HP).

## 4. Discussion

In this multimodal MRI study of long-term neurological sequelae among COVID-19 survivors, we leveraged structural, diffusion, functional, and per-fusion imaging to study alterations in the brain. By comparing 76 CRPs with 51 HCs, and further classifying CRPs into hospitalized (HP; N = 21) and non-hospitalized (NHP; N = 46) subgroups, we delineated neurological effects in CRPs and how infection severity modulates microstructural and functional connectivity. Below, we integrate our findings with existing literature, explore plausible mechanistic underpinnings, acknowledge study limitations, and outline future directions.

### 4.1. Regional Specificity of Structural Changes

We conducted a detailed analysis of the morphological changes in the brain as a long-term effect of the COVID-19 infection. We started by comparing TIV, GMV, and WMV between the HCs and CRPs to test for any signs of widespread cortical atrophy. We observed that the TIV, GMV, and WMV did not differ between CRPs and HCs. Instead, subcortical analysis revealed selective volume reductions in right-hemisphere basal ganglia (putamen, caudate, pallidum) and limbic nodes (thalamus, hippocampus, amygdala). These findings align with Hiene *et al*. who similarly observed basal ganglia atrophy in the COVID-19 recovered patients [35]. Structural damage in the limbic system has also been consistently observed in COVID-19 survivors, ranging from GM loss in the thalamus [36] to changes in the orbitofrontal cortex [11], hippocampal and parahippocampal regions [37, 9, 11]. The absence of global changes alongside focal subcortical loss suggests that SARS-CoV-2 may preferentially target deep grey nuclei, perhaps via microvascular injury or direct neurotropism, rather than inducing diffuse neu-rodegeneration. Surface-based morphometry further localized cortical thinning and volume loss to paralimbic regions: the medial prefrontal cortex, insula, and posterior superior temporal gyrus. These loci have been implicated in attentional control, salience detection, and reward processing; functions commonly reported as impaired in “brain fog” and fatigue (lack of motivation) [2, 38]. Our results extend prior studies [6, 8] by demonstrating that even subtle cortical alterations are reproducible across cohorts and imaging platforms.

### 4.2. Microstructural Integrity of Limbic White Matter

The dMRI analysis revealed that UF exhibited decreased FA and increased RD in CRPs relative to HCs, with greater deviations in the HP sub-group. These changes are consistent with axonal injury and edema in these regions. The UF interconnects orbitofrontal and anterior temporal regions, supporting emotional regulation and memory retrieval [39, 40]. Concurrent abnormalities in cingulum bundle subdivisions (CC, CH) further suggest a systemic disruption of limbic circuitry. These results are the extension of the outcomes reported in our previous study [34]. These patterns of damage to limbic white matter were also observed by Diez-Cirarda *et al*. [9], Paolini *et al*. [11], and Kremer *et al*. [41], who reported elevated MD and altered AD in the cingulum of post-COVID individuals. The combination of increased FA and lowered RD is consistent with extracellular fluid accumulation (edema), potentially reflecting stressed intra-axonal space or a microvascular leak in this tract. Overall, these observations indicate system-wide damage in the connectivity of the limbic system.

### 4.3. Functional Connectivity Alterations with Severity

Resting-state ICA did not detect cohort-wide network differences, but stratification by severity uncovered that HPs exhibited significantly reduced connectivity between the default mode network (DMN-A) and insular cortex, as well as between DMN components and the caudate nucleus. The insula–DMN coupling underpins the ability to flexibly shift between internally and externally focused attention [42]; its disruption may therefore underlie deficits in sustained attention and cognitive flexibility. Reduced DMN–caudate connectivity also correlates with mood dysregulation, anhedonia in depression [43], and sleep disturbances [44], offering a potential link to the lack of motivation, insomnia, and fatigue reported by many survivors. Our findings build on Leitner *et al*. [10] and Voruz *et al*. [12] by showing that the severity of the acute infection predicts enduring functional network impairments.

### 4.4. Perfusion Imaging and Vascular Changes

Despite previous reports of CBF reductions in post-COVID cohorts [13, 14], our ASL analysis across 68 ROIs found no statistically significant group or severity effects after FDR correction. The strongest sub-threshold trend in the left pallidum may warrant targeted follow-up, but overall, these results suggest that macroscopic perfusion deficits, if present, may be transient or heterogeneous across individuals. Alternatively, our cross-sectional study, averaged at 3-6 months post-infection, may have missed early perfusion abnormalities that normalize over time.

### 4.5. Mechanistic Implications

Collectively, our findings support a model in which SARS-CoV-2 induces focal neuronal and axonal injury within deep grey nuclei and limbic networks, likely mediated by endothelial dysfunction, inflammatory cytokines, and possible neuroinvasion [45]. Here, we report localised atrophy in basal ganglia and paralimbic cortices, compromised integrity of the uncinate fasciculus and cingulum bundle, and severity-dependent DMN decoupling in COVID survivors. All together, these imaging alterations correspond closely to their self-reported symptoms. A majority of CRPs (40/59) reported persistent fatigue, and 26/59 noted attentional lapses weeks after recovery. The focal damage to the deep grey nuclei of the basal ganglia and the insular cortex outlines abnormalities in the reward processing circuitry in CRPs, aligning with reports of loss of motivation, and cognitive fatigue [4, 46].

Furthermore, altered FC between the DMN and the insular cortex reflects disruptions in the ventral attention and salience networks in the CRPs, providing an anatomical substrate for complaints regarding “brain fog”. Atrophy in the subcortical (thalamus, amygdala, hippocampus) and cortical (prefrontal cortex, anterior cingulate cortex) limbic nodes, coupled with microstructural damage to the UF and cingulum bundle, may underlie the high prevalence of memory impairment in CRPs. We also observed structural and functional disruptions in the caudate nucleus, which is known to facilitate slow-wave Rapid Eye Movement (REM) sleep [44]. These aberrations could provide a possible explanation for reports of unrefreshing sleep (27/59) in CRPs. Overall, this concordance between neuroanatomical findings and behavioural reports reinforces a unified model whereby SARS-CoV-2 preferentially injures limbic–basal ganglia circuits, yielding the multidomain symptomatology of post-COVID syndrome.

## 5. Limitations

Certain limitations of this study need to be acknowledged to contextualize the findings and inform future research directions. One key limitation is the absence of comprehensive pre-infection medical histories and detailed comorbidity data for both CRPs and HCs. Pre-existing conditions such as hypertension, diabetes, or respiratory illnesses, known to influence neurological health, could have contributed to the observed differences in brain structure and connectivity. While individuals with a history of neurological disorders or brain trauma were excluded to minimize confounding factors, subtle baseline health differences may still have influenced the results. Future studies incorporating pre-infection medical records, additional control groups with other viral illnesses, and longitudinal follow-ups will be essential to disentangle the effects of COVID-19 from underlying health conditions.

Another potential limitation is the risk of undetected asymptomatic subjects with COVID-19 infections within the HC group. Given the high prevalence of asymptomatic cases during the pandemic and the limitations of testing at the time, it is possible that some control subjects had prior, unconfirmed SARS-CoV-2 infections. Although rigorous screening criteria were applied, including negative RT-PCR confirmation for symptomatic individuals, asymptomatic infections could have reduced the contrast between groups. Future studies employing serological testing or antibody assays may help better classify subjects and refine group comparisons.

## 6. Conclusion

This multimodal MRI study of 76 COVID-19 survivors and 51 matched controls reveals that long-term neurological sequelae concentrate in inter-connected limbic–basal ganglia circuits rather than manifesting as diffuse brain injury. We observed focal subcortical atrophy (thalamus, hippocampus, amygdala, putamen, caudate, pallidum), paralimbic cortical thinning, and microstructural disruption of the uncinate fasciculus and cingulum bundle. Resting-state analyses showed severity-dependent decoupling between default mode and salience networks, while perfusion deficits normalized by 3–6 months post-infection. Crucially, these circuit-specific alterations map directly onto patients’ persistent fatigue, memory impairments, attentional lapses, and sleep disturbances, supporting a unified model of Post-COVID brain injury, likely driven by endothelial dysfunction and neuroinflammation.

Our findings establish precise imaging biomarkers for the post-COVID syndrome and underscore the need for targeted interventions, such as neural stimulation of the salience network or dopamine-enhancing therapies, to restore circuit function. Future longitudinal work integrating high-field MRI, PET microglial markers, and fluid biomarkers will be essential to chart recovery trajectories and optimize personalized rehabilitation strategies.

## Data Availability Statement

All data produced in the present study are available upon reasonable request to the authors.

